# ‘I don’t know anybody who said, “oh great, let’s get measles”’: a qualitative study of responses to childhood vaccinations (MMRV) among Orthodox Jews in Jerusalem following the 2018-19 measles outbreaks

**DOI:** 10.1101/2022.01.04.22268713

**Authors:** Ben Kasstan

**Author notes:** Corresponding author at: Centre for Health, Law & Society, University of Bristol Law School, Bristol, UK BS8 1RJ.

## Abstract

Measles outbreaks have emerged among religious minorities in the global north, which cross regional and national boundaries and raise implications for measles elimination targets. Yet, studies are ambiguous about the reasons that underlie non-vaccination in religious populations, and whether and how religious “beliefs” influence vaccine decision-making among populations with suboptimal vaccination coverage. In 2018-19, Israel experienced the largest measles outbreaks in a quarter century – the burden of which disproportionately affected Orthodox Jewish neighbourhoods in Jerusalem. The objective of this study was to explore how Orthodox Jewish households in Jerusalem responded to the measles outbreaks in their neighbourhoods and how they viewed childhood vaccination (MMRV) during a public health emergency.

Research methods primarily consisted of 25 in-depth semi-structured interviews conducted with 23 household heads, and 2 public health professionals involved in planning and implementation of vaccination services. Thematic analysis generated five key themes, i) where the issue of sub-optimal vaccination uptake was perceived to be located; ii) how responsive people and services were to the measles outbreaks; iii) the sources of information used in vaccine decisions by religious parents; vi) whether vaccination was deemed a religious issue; and v) how vaccination influenced social relations within religious neighbourhoods.

Results demonstrate parental investment in protecting child health, with decisions around vaccination reflecting vaccine efficacy and safety, and the risk of measles transmission. Household heads across all Orthodox Jewish backgrounds were not apathetic towards measles transmission. No religious “beliefs” were identified for non-vaccination among the household heads in this cohort. Rather than relegating suboptimal vaccination uptake among religious minorities and populations as an issue of religious “beliefs,” quality social science research should examine – and clearly convey – how religion influences vaccine decision-making. Such clarity can help to avoid stigmatizing religious minorities and populations, and to plan for appropriate vaccination programmes and promotion campaigns.

## Introduction

Routine childhood vaccinations are available free of charge from maternity and infant care clinics in Israel, which are known as *tipot halav* (“drops of milk,” pl.). Israel has a long-standing two-dose measles (MMRV) vaccination plan that has led to overall high coverage.^1^ Yet, coverage is consistently lower in Orthodox Jewish neighbourhoods in Jerusalem^2,3^ and central Israel,^4^ which has led to a series of localized and national measles outbreaks.^2-6^ The largest measles outbreaks in a quarter century were recorded in Israel and the USA in 2018-19, which were linked to unvaccinated Orthodox Jewish travellers.^7,8^ In Israel, the burden of the 2018-19 outbreaks were disproportionately located in Jerusalem – where 52.9% of measles cases occurred.^5^ Most studies since the 2018-19 measles outbreaks have been epidemiological, highlighting that Orthodox Jewish populations were underimmunized and which public health interventions were developed to boost vaccination coverage levels.^2-5^ The aim of this study is to explore responses to the measles outbreaks among Orthodox Jews and how they viewed childhood vaccination (MMRV) during a public health emergency.

### Background: Religion, Vaccination, and Influences on Decision-Making

From Orthodox Protestants and Jews to Amish minorities, suboptimal childhood vaccination coverage among religious minorities and movements in the global north is an increasing public health challenge.^8-10^ Religious minorities are not isolated populations, and suboptimal childhood vaccination coverage levels have led to persistent outbreaks of preventable disease that cross regional, national and international boundaries.^7,11^ Emerging evidence suggests that COVID-19 vaccination uptake and coverage is suboptimal among religious and ethnic minorities,^12^ indicating that the issue is not confined to child health.

Yet, public health and social science research remains ambiguous about *how* religion or ‘beliefs’ influence vaccine decision-making. Immunization managers (who maintain responsibility at national levels for implementing immunization programmes) often cite ‘religious beliefs’ as a causal factor of vaccine hesitancy and low uptake.^13^ Social scientists instead argue that how religious ‘beliefs’ constitute a reason *not* to vaccinate is rarely ever explained.^14^ No major religious doctrine opposes vaccination.^15^ Religious legal issues that affect uptake of individual vaccines (e.g. influenza vaccines produced with porcine^16^) do not explain suboptimal engagement with routine vaccination programmes. Jewish law (*halachah*) does not explicitly endorse or prohibit vaccination,^17^ but is widely interpreted by rabbinic authorities as an important way to protect child health.^18^

Beyond doctrine, the influences on vaccine decision-making that can be considered ‘religious’ include consultation with religious authorities and social networks, and personal assessments of religious texts and laws. Religious authorities are often viewed as central to promoting confidence in vaccination programmes,^19,20^ but rabbis do not appear to be routinely consulted by Orthodox Jews in vaccine decision-making.^21,22^ Attention to non-vaccination activism has highlighted the strategies that activists use to promote their cause and specifically to target self-protective religious minorities.^23^ Scholars also note that pro-vaccine activism takes on situated (context-specific) forms, and illustrates how examining public support *for* vaccination is helpful to illustrate the kinds of impact that are sought in relation to local context.^24^

Orthodox Jewish minorities in Israel, Europe and North America have experienced recurring measles outbreaks and share a common pattern of sub-optimal vaccination coverage.^8,25,26^ Past studies have identified a gap between higher primary care consulting rates but lower levels of vaccination uptake among Orthodox Jews in the UK.^27^ Barriers to vaccination uptake include inflexible services,^25^ particularly because of larger family sizes. Additional influences on uptake include misinformation,^23,28^ and, in Israel, a reluctance to engage with public health services.^6^ Studies exploring non-vaccination among Orthodox Jewish minorities note that perception of risk and safety are primary influences on vaccine decisions.^21^

This paper asks how responsive are underimmunized minority populations to measles outbreaks? How are decisions around vaccination made in religious families? And what lessons emerge from the outbreaks for public health delivery strategies? The present study addresses these questions by drawing on a qualitative study conducted with Orthodox Jewish families during the 2018-19 measles outbreaks. Against the epidemiological backdrop of suboptimal vaccination coverage among Orthodox Jewish minorities in Israel, North America and Europe, the paper focuses on the perspectives of migrant families in Jerusalem to understand what informs vaccine decision-making in diasporic networks. Implications are raised beyond the case at hand for social scientists, who ambiguously cite religion or ‘beliefs’ as impacting delivery of vaccination programmes.

## Methods

To examine responses to suboptimal vaccination coverage among religious minorities affected by the 2018-19 measles outbreaks, a qualitative study was conducted in Jerusalem and the surrounding region between October 2019 and March 2020. Research methods consisted of: i) 25 in-depth semi-structured interviews conducted with household heads (HH) (21 mothers; 2 fathers) and 2 public health professionals (PH) involved in planning and implementation of vaccination services; ii) ethnographic research (participant observation) and informal conversations were conducted in non-vaccination and pro-vaccination advocacy events, study halls, and religious book stores (which offer a plethora of information pertaining to family health)^29^; and iii) discourse analysis of measles outbreaks in international and local Jewish media.

Participants were recruited through past fieldwork connections, and snowball sampling. Interviews took place, typically, in people’s homes amidst their family routines, but also public spaces if this was more comfortable (usually for women) and learning halls (for men). Three key participants provided repeat interviews. The study design included parents with a range of vaccine positions; parents accepted vaccinations according to recommended guidelines, whereas others had either accepted vaccines selectively, delayed uptake, or refused to vaccinate their children altogether. Participants had family sizes ranging from two to seven children. Analysis of the data was inductive and thematic, whereby theoretical insights emerge from prolonged engagement with the data rather than being pre-conceived.^30,31^

Ethnographic fieldwork ended prematurely due to restrictions on public movement imposed by the Israeli Ministry of Health amidst the COVID-19 pandemic in March 2020. Interviews were recorded using a digital audio recording device, when permission was granted, and detailed notes recorded. Recordings from interviews and participant observations in the field were transcribed verbatim, and analysed thematically. Participants provided written consent. Approval to conduct this study was obtained from the Ethics Committee of the Faculty of Social Sciences of the Hebrew University of Jerusalem on 22 December 2019.

### Place, population and particulars

Jewish Orthodoxy consists of a diverse and broad continuum, and groups can be distinguished in terms of origin and stringencies in which religious law is interpreted and practiced. Jewish Orthodoxy includes modern Orthodox, Orthodox, religious Zionist (*Dati Leumi)*, and Haredi Jews. Whereas modern Orthodox and Orthodox Jews attempt to reconcile religious piety with national curricula and professional opportunities, Haredi Jews can be distinguished by their self-protective stance and controlled exposure to non-religious education and training.

Most recent social science research focuses on sub-optimal vaccination coverage among Haredi Jews,^21,22^ reflecting epidemiological studies that locate outbreaks among Haredi Jewish neighbourhoods. Denominational backgrounds are not routinely recorded in health records in Israel, and cases of measles among Haredi Jews are estimated according to geographic locations.^5^ The population of Israel is approximately 9 million, 74% of which is Jewish.^5^ The Haredi Jewish population constitutes approximately 1 million, or 12% of the Israeli population.^31^ High fertility rates are a feature of Haredi Jewish families, and about 16% of the Haredi population is under the age of 16.^31^

Studies tend to approach Orthodox Jewish minorities as separate analytical categories, though looking across the groups can yield insights into shared challenges around family health.^32^ I worked across these communal divisions to discern the shared and situated influences on vaccine decision-making (Figure 1). Interview participants were Orthodox Jewish parents from all of the above-named groups. Participants originated from the USA (17 out of 23), and to a lesser extent Australia, Canada, South Africa, and the UK. The reason for focusing on parents with migrant backgrounds was because the 2018-19 international measles outbreaks were, as mentioned, linked to unvaccinated Orthodox Jews travelling between Israel and the USA,^7^ and because common issues of suboptimal vaccination coverage have been recorded in Israel, North America and Europe.

**Figure 1:**
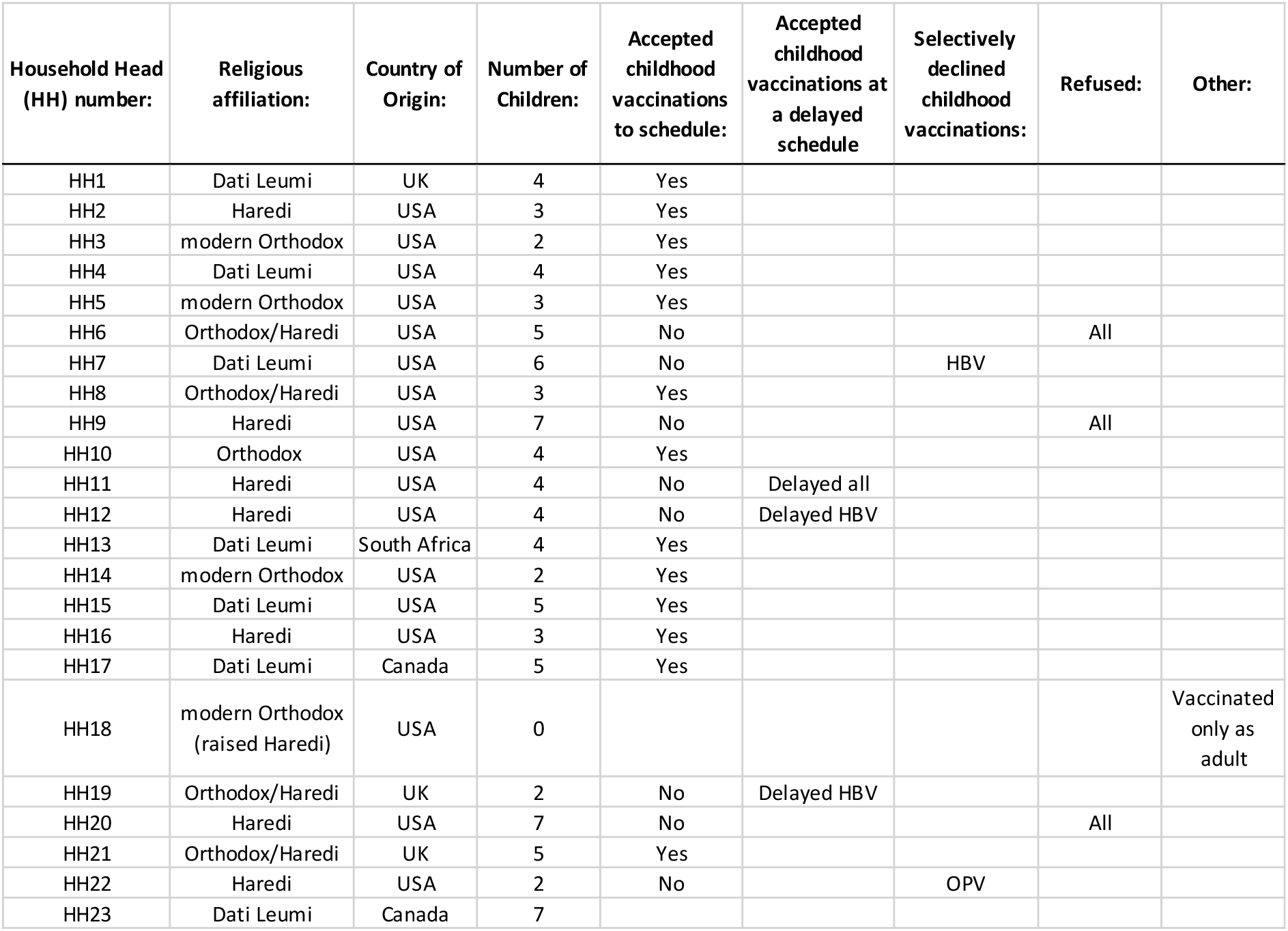
Religious affiliations of participants and vaccination decisions.

## Results

Analysis of results raised five key themes, i) where the issue of sub-optimal vaccination uptake was perceived to be located; ii) how responsive people and services were to the measles outbreaks; iii) the sources of information used in vaccine decisions by religious parents; vi) whether vaccination was deemed a religious issue; and v) how vaccination influenced social relations with Orthodox Jewish minorities.

### i) Where did religious minorities locate the issue of sub-optimal vaccination uptake?

HHs were aware that the 2018-19 measles outbreaks had been centred in Orthodox Jewish neighbourhoods. HHs associated suboptimal MMRV vaccination uptake with Haredi communities – primarily due to larger family-sizes and subsequent issues of convenience:

‘They say they’re not vaccinating in the Haredi world, but I think it’s a scheduling thing. You have a bunch of kids, you just don’t want to go to *Tipat Halav*, you want your baby to sleep, “so maybe I’ll do it later.”‘ (HH6)

Haredi HHs, however, were keen to convey their acceptance of childhood vaccinations despite the public health focus of suboptimal vaccination coverage among Haredi neighbourhoods: ‘Everybody I know gives shots to their kids and if they don’t give, they’re a little lazy and they give a month later. That’s like the worst you get. There’s small groups, it’s true, in Meah Sha’arim [Haredi neighbourhood in Jerusalem], they don’t vaccinate.’ (HH12)

Blaming religious minorities for suboptimal vaccination coverage was considered unfair and accusatory, implying that Haredi parents did not value the role of vaccinations in protecting child health:

‘Why people like to blame the Haredi community, specifically, is because we’re generally insular and “we refuse to assimilate” or because “we don’t believe in science.” The thing specifically with vaccines, I never saw it as an issue. It’s just like taking antibiotics if you have strep. That’s what you do.’ (HH22)

A public health professional noted that the issue of underimmunization was not only an issue with Haredi Jewish minorities:

‘The focus on Haredi Jews ignores the broader range of groups with vaccine hesitancy, the *Dati Leumi* are not addressed in the literature. Most are like the general population and do vaccinate, there is a small group that don’t. I feel they are the future problem, if they’re not dealt with, they will be.’ (PH1)

While public health and social science studies tend to locate suboptimal vaccination coverage among Haredi neighbourhoods, PH professionals were concerned that the issue of ‘vaccine hesitancy’ spanned the full continuum of Orthodox Jewish sectors in Jerusalem. HHs who identified as modern Orthodox, Orthodox and *Dati Leumi* did deliberate over childhood immunisations. However, they were less concerned with the MMRV and more with hepatitis B (HBV), as the first dose is administered on the day of birth in Israel. HHs deliberated over the timing of HBV, and a minority (2) of participants in the sample took the decision to delay administering HBV to 1 month:

‘There is a vaccination they get when they are born, the day they’re born, hep B. I delayed that, I didn’t want a newborn to go through that and I’m glad I did.’ (HH19)

Just 1 HH declined the HBV, perceiving it to be unnecessary due to the association between hepatitis B with sexual transmission:

‘I did the others, only hepatitis B and I guess it was just a newborn, and that hepatitis B is sexually transmitted and it’s not really necessary or whatever.’ (HH7)

Decisions to delay or refuse childhood vaccinations were not associated with religion, but rather what vaccines are perceived as necessary to protect child health.

### ii) How responsive were people and services to the measles outbreaks emerging in Orthodox Jewish minorities?

The severity of the 2018-19 measles outbreaks brought a sense of urgency to receive vaccination, including for unvaccinated adults:

‘There was an outbreak of measles a year ago and we sat down as a family and we spoke about it. I said, “I’m not vaccinated, what are we supposed to do?” My mum really encouraged it, actually, which is funny because she was the one who discouraged it. All our life she didn’t want to do it and all of a sudden she had said, “there’s a measles outbreak, you need to go and get vaccinated.”‘ (HH18)

HH perceived co-religionists to be responsive, albeit reactively, to transmission risk during the measles outbreaks in their neighbourhoods. Exposing children to measles was not considered to be desirable:

‘I was in the *Tipat Halav* with my two-year old when she was an infant and it was the outbreak. I remember hysterical parents coming in because they were at a wedding, or a *bar mitzvah*, and there was someone there who afterwards turned out to be sick and they were hysterical. I’m talking about a religious neighbourhood. I don’t know anybody who said, “oh great, let’s get measles.” That is not a thing, for sure.’ (HH12)

A small minority of HHs (1) wanted to expose children to measles during the outbreaks on the basis that ‘natural immunity’ (exposure via infection) was viewed as more effective than vaccine-induced immunity:

‘I was hoping he [child] would get measles. I tried to get it from one family, I found who got it, they didn’t want to give it. They were having a bad experience with their children and they didn’t feel it was right to pass it on. They had not vaccinated their kids, so they got measles.’ (HH6)

Yet, vaccination services were not perceived to be as conducive as possible for parents who had made appointments for their children to receive the MMRV during the outbreaks:

‘The *Tipat H*alav had reduced hours and coverage at the time of the measles outbreak and the appointment was moved to a branch that was totally not accessible for me. I don’t have a car. It was really in a not convenient location. They didn’t cancel, but to me that’s like cancelling. I waited a year for this vaccine, *it will not wait one more day*.*’* (HH5, her emphasis)

HH emphasized that services were not proactive in planning for the higher fertility rates in Haredi neighbourhoods, which places a subsequent pressure on maternity and infant clinics:

‘The *Tipat Halav* is like an issue. It’s hard to get through to them at times, to make an appointment. It’s very busy. They run late. So, it’s hard to reach them. I live in a place with high birth rates, so from a public health perspective, it’s *really* bad.’ (HH22)

### iii) Which sources of information were used to inform vaccine decision-making?

Parents cited diverse strategies to inform their decisions on vaccination, including information produced by Israeli healthcare services and their countries of origin. One parent emphasised how they followed the advice of their local *Tipat Halav*:

‘I mean it was whatever they said in *Tipat Halav*, I went ahead and did, and there was never any question of not doing it, or doing anything different to what they recommended.’ (HH1)

Parents who had settled in the country more recently continued to use guidance informed by the Centre for Disease Control (CDC) when vaccinating their children in Israel:

‘I just followed the CDC schedule with my daughter.’ (HH2)

Yet, Haredi HH who were cautious of vaccination questioned guidance from bodies such as the CDC that presented childhood vaccinations as safe and effective and sought out further information about the risk of side-effects:

‘I’ve tried to go to the CDC website, and they’ll say, ‘vaccines are safe.’ Now it can’t be that simple, so when they sound so condensing and simplistic, that makes me nervous. I think in the Orthodox world, we’re willing to question authorities more.’ (HH16)

The Internet was used in combination with medical opinion, to help generate questions for encounters with trusted healthcare professionals:

‘If I’m curious about something, then I’ll Google it, or if I want to know what questions to ask, I would Google it. But in terms of getting information that I feel is trustworthy and reliable, it would come from someone with whom I have a direct relationship of trust with – which would be my physician.’ (HH10)

Medical opinion on childhood vaccinations was also viewed as one of several information sources to consider:

‘Rather than “I’m going to listen to what my doctor says,” a doctor is just one opinion amongst many that I’m going to take into account.’ (HH2)

Medical opinion on vaccination was rarely balanced against the opinions of religious authorities:

‘When it came to these types of decisions, I didn’t see any need at all to go to a rabbi, because it was very clear, the science wasn’t in any shape or form against our religious principles. For this type of issue, it would never have occurred to me to go to a rabbi.’ (HH22)

A minority of HHs (2) approached both physicians and religious authorities for support with vaccine decisions, especially if partners held opposing positions. A parent of seven children recalled how he was advised not to vaccinate by a physician in the USA, but to listen to his physician by a rabbinic authority:

‘The paediatrician said the fact that you have two kids that have special needs gives me some concern of what’s going on, if I was you, I would not vaccinate. Then when I talked to the rabbi, he said, ‘listen to your doctor.” So that put me in a little bit of a bind. I asked a rabbi two weeks ago, “should we vaccinate or not?” He said, “don’t touch it” – because my wife is against it, and I’m not, so he doesn’t want to get involved.’ (HH23)

Hence, disagreements over childhood vaccination emerged within HHs.

### iv) Was vaccination viewed as a religious issue?

HHs who accepted vaccines did not view childhood vaccination as a religious issue:

‘As far as I can see, never has there ever been a [rabbinic] statement saying, “it’s prohibited to,” or “one should not vaccinate [children].” So, it never was a religious issue in that way.’ (HH15)

Common to parents who accepted vaccinations was the perception that preserving health was required by Jewish law, which was interpreted expansively to include vaccination:

‘you’re required to do whatever is going to protect your health, in other words, ‘*v’nishmartem meod le’nafshotechem*.’ (HH11)

While parents unanimously agreed that preserving health (*pikuach nefesh)* is crucial from the standpoint of Jewish law, the issue of vaccination raised conflicting pursuits of *how* to protect child health. Parents who refused vaccination based on safety and risk calculations noted that vaccination did not constitute a religious legal issue. Yet, they interpreted religious teachings around health protection to suit their positions against vaccination:

‘I never thought that it was a religious issue, I thought it was a personal, faith-formed decision. Faith as opposed to *halachah*. If you can show me in the Torah [Hebrew Bible] where it says, “you must vaccinate your child,” then I would have to vaccinate my child, but it says, “*v’nishmartem meod le’nafshotechem*,” that means, “that you should be very careful to take care of the body that I gave you, very careful, *nishmartem maod*.” That’s up for interpretation.’ (HH20)

Hence, parents who accepted and refused vaccinations used the same religious teachings to underscore their decisions.

### v) Did vaccine decisions influence social relations in religious minorities?

HHs who accepted childhood vaccinations were concerned about transmission risk in Orthodox Jewish neighbourhoods during the 2018-19 measles outbreaks. HHs (x3) were especially anxious that unvaccinated infants under the age of 12 months would be exposed to measles, and actively took steps to exclude unvaccinated children from social events. A mother gave birth to a daughter amidst the 2019 measles outbreaks explicitly sought to exclude non-vaccinated children from a naming ceremony (*Simchat Bat*) by writing on the invitations (in English):

‘as there will be little immune systems at the event, please do not attend if you are not feeling well or have not been vaccinated.’ (HH3)

She did not specify on the invitations which vaccine-preventable diseases she was concerned about, and more broadly parents had taken steps to wholesale exclude non-vaccinating families from religious and communal spaces such as synagogues. Concerned about the risk of transmission, participants took steps to lobby the religious authorities of their local synagogues to exclude non-vaccinating families – which were not always successful:

‘So, when my youngest who is not yet one [age], and the measles stuff broke, I got really really anxious, right, ‘coz she’s not vaccinated. I got in touch with the *Gabbai* [synagogue warden] and I said, “what’s the deal? Are there unvaccinated people in our community?” And he was very, very, ambiguous. It made me very uncomfortable. Basically I said, “I want you to make an announcement that unvaccinated people are not welcome.” He said, “no, we can’t do that, we can’t exclude people.” I said, “of course you can. If you won’t exclude them, you’re excluding me.”‘ (HH5)

Other HHs were successful in lobbying religious authorities to request vaccination status as part of synagogue membership due to the urgency of the measles outbreaks:

‘When the measles outbreak took place, my husband spoke to the *Gabbai*, and they discussed it with the *Rav* [rabbi] and they said very quickly, “we’ve never asked about it before because we didn’t see it as something imminent, but now you’re right.” They posted in the community WhatsApp group that Jerusalem had some of the largest outbreaks, there was a baby who died last year and there were two older individuals who died last year [from measles]. They said, “we believe that this is important, if you feel otherwise, that’s your decision, but we ask that you don’t come to *shul* [Yiddish, synagogue].”‘ (HH8)

More broadly, HHs described conflicts with extended family members over the responsibility to vaccinate because of the measles outbreaks, which became aired using social media and telephone applications:

‘In the family WhatSapp group, there was a back and forth about the *halachic* obligation to vaccinate and the people that didn’t vaccinate felt persecuted and they left the WhatsApp group. It became a big family to-do.’ (HH4)

HHs that opposed vaccinations rejected claims that they constituted threats to public health:

‘There’s the people – like one of my siblings – who said that we’re selfish, ‘you don’t care about anybody else,’ and ‘terrorists’ has been thrown out last year with the measles [outbreaks]. It’s very painful, as a religious person.’ (HH20)

Hence, the decision to refuse childhood vaccinations carried a social sanction within and between Orthodox Jewish social networks.

## Discussion

While overall MMRV coverage in Israel is high and estimated to be 97% and 96% for the first and second doses respectively, unprecedented measles outbreaks emerged in 2018-19.^5^ Underimmunization was widespread among measles patients – most of whom (75%) were children.^5^ Epidemiological and social science studies have located the issue of suboptimal vaccination coverage and measles outbreaks among Haredi Jewish neighbourhoods.^2-6,21,22^ In this study, PH professionals were concerned by a deficit of research examining vaccine decision-making across Orthodox Jewish sectors in Jerusalem and Israel.

The study uncovered diverse responses to the issue of suboptimal vaccination coverage within a range of Orthodox Jewish minorities in Jerusalem. Parents were aware of, and responsive to, the measles outbreaks that emerged in Orthodox Jewish minorities and sought to secure MMRV for their children and manage communal spaces to avoid measles transmission. A minority of parents that opposed childhood vaccination perceived acquired immunity through infection to be more protective than immunity acquired through vaccination, reflecting research conducted in non-vaccinating populations more broadly.^34^ Routine vaccination services were not viewed as flexible or conducive to accessing vaccinations during the outbreaks, including in religious neighbourhoods that are characterised by large family sizes. Parents that accepted and refused childhood vaccinations made decisions primarily based on evaluations of vaccine safety and transmission risk.

Evidence suggests that there is a transnational pattern of suboptimal vaccination coverage leading to measles outbreaks among Orthodox Jewish minorities in Israel, North America and Europe.^7,8,11,25^ This study focused on migrant Jewish households to explore the influences on decision-making in diasporic networks. Mainstream public health information, produced in Israel and abroad, was accessed among all Orthodox Jewish sectors in this sample. Religion or religious law was not viewed as the primary issue affecting suboptimal vaccination uptake across all Orthodox Jewish sectors represented in the sample. Religious authorities were rarely consulted as part of vaccine decision-making, and when approached, would defer to the advice of medical professionals. At other times, rabbinic authorities would prefer not to interfere if parents held opposing views on childhood vaccination. Co-religionists were expected to conform with MMRV acceptance, with allowances made for accepting vaccination at a delayed schedule – though refusal was socially sanctioned.

Studies note that immunization managers often cite “religious reasons” or “beliefs” as a causal factor for underimmunization, without specifying what such reasons involve.^13^ Quality social science research should examine – and clearly convey – how religion or ‘beliefs’ influences vaccine decision-making to avoid stigmatizing religious minorities, and to plan for appropriate vaccination programmes and promotion campaigns. The results of this study offers indicators for public health and social science scholarship to gage the influence of religion on vaccine decision-making (Figure 2). Indicators of religion on vaccine decision-making will always vary according to context.

**Figure 2:**
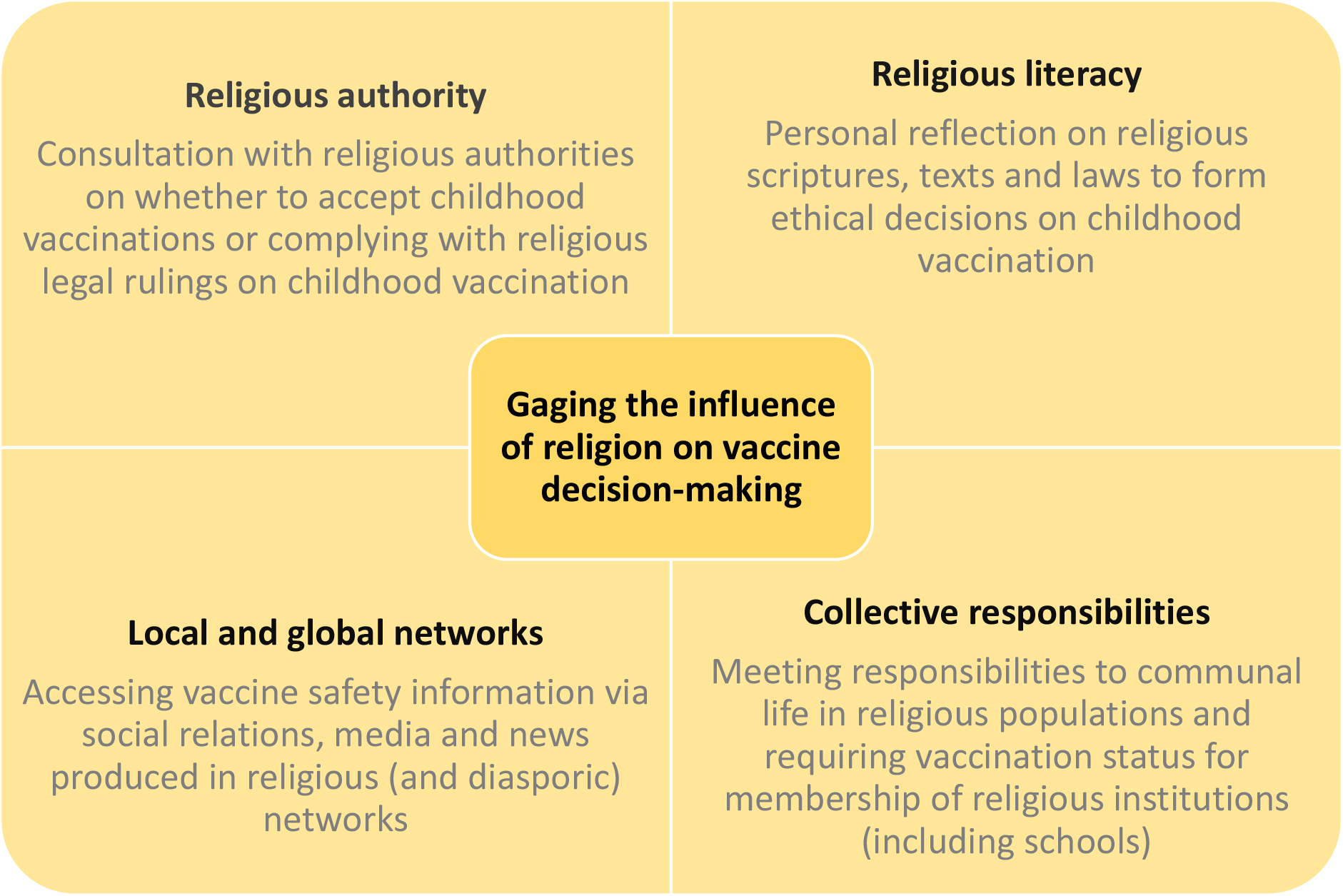
Indicators for public health and social science scholarship to gage the influence of religion on vaccine decision-making.

Public health studies conducted among Haredi Jews in Jerusalem during the 2018-19 measles outbreaks report ‘remarkable population compliance’ with vaccination programmes.^2^ In England, public health agencies and a Jewish rapid response service collaborated to co-deliver COVID-19 vaccinations for Haredi Jews, suggesting that convenience can impact uptake.^35^ This body of evidence indicates that broad-scale or organized anti-vaccination sentiments is not the major issue that underlies underimmunization in this population. Yet, participants in this study indicated that Orthodox Jewish populations and vaccination services appear to be *reactive* to measles outbreaks. The need for flexible and accessible service delivery raise implications for designing vaccination services that *proactively* address coverage in religious minority contexts that are characterized by high fertility.

Most studies of childhood vaccine decision-making among religious minorities are conducted in the global south, especially to review responses to top-down and mass polio vaccination programmes. Social scientists note that refusal among religious minorities reflects concerns of safety or mistrust in public health and government services, and a perception that quotidian threats to child health (malaria, sanitation) are overlooked in the focus on polio eradication.^36,37^ The resurgence of measles and vaccine-preventable diseases (VPD) in North America, ^7,11^ Europe^25^ and Israel^8^ raise practical and theoretical implications for how elimination of VPDs is approached in settings that are characterised by different public health challenges and resources, and the influence of diasporic networks. Further research is required to understand responses to vaccination amidst the resurgence of VPDs and failure to reach elimination targets.

Measles elimination in the World Health Organization (WHO) European Region is undermined by underimmunization in childhood and ‘populations at risk’ from suboptimal coverage.^38^ The World Health Organization ranked ‘vaccine hesitancy’ as among the top ten threats to global health in 2019, commensurate with the dangers posed by antimicrobial resistance and climate change.^39,40^ Media coverage of the COVID-19 pandemic have framed ‘vaccine hesitancy,’ misinformation and refusal as major issues affecting Orthodox Jewish populations in Israel and the US.^41^ Yet, as has been noted in the context of declining vaccination coverage in England, the public health community should not contribute to making the challenge of anti-vaccination advocacy bigger than it actually is – and to instead focus on maximizing protection against VPDs by ensuring that immunization programmes are implemented as effectively as possible.^42^ Similarly, religious “beliefs” appear to be classed as a distinct form of hesitancy and refusal among religious minorities, when influences on vaccine decision-making and uptake concur strongly with broader populations. Over-emphasising the problem of religious ‘refusal’ or resistance only hinders the design of appropriate solutions and shared goal of health protection.

### Strengths and Limitations

Clarifying the role of religion in vaccine decision-making is a key strength of this paper, as public health studies reference ‘religion’ and ‘beliefs’ ambiguously when discussing influences on vaccination uptake. To my knowledge, this is the first qualitative study exploring responses to childhood vaccination among diverse Orthodox Jewish sectors. The qualitative methodology of this paper enabled a comparison of influences among Jewish Orthodox minorities, but the modest sample size constitutes a limitation on extrapolating results more broadly.

## Conclusion

The 2018-19 measles epidemics disproportionately affected Orthodox Jewish neighbourhoods in Jerusalem. Epidemiological studies have highlighted that Orthodox Jewish populations were underimmunized and what public health interventions were developed to address the outbreaks. This qualitative study has contextualised how Orthodox Jews in Jerusalem responded to the measles outbreaks in their neighbourhoods and examined how childhood vaccination (MMRV) is viewed as an important part of protecting and promoting child health. Public health studies should clearly stipulate how religious ‘beliefs’ cause suboptimal childhood vaccination coverage to avoid stigmatizing minorities and populations and to appropriately design promotion campaigns.

## Data Availability

The data produced in the present study is confidential as a condition of participant consent.

## Ethics

Approval to conduct this study was obtained from the Ethics Committee of the Faculty of Social Sciences of the Hebrew University of Jerusalem on 22 December 2019

## Acknowledgements

Thanks to all participants for sharing their time, Ami Dabush, and Nurit Stadler.

